# Development and calibration of a mathematical model of HIV outcomes among Rwandan adults: informing equitable achievement of targets in Rwanda

**DOI:** 10.1101/2024.09.06.24313223

**Authors:** April D. Kimmel, Zhongzhe Pan, Ellen Brazier, Gad Murenzi, Benjamin Muhoza, Marcel Yotebieng, Kathryn Anastos, Denis Nash, the Central Africa International epidemiology Databases to Evaluate AIDS (CA-IeDEA)

## Abstract

**Background:** We developed and calibrated the Central Africa-International epidemiology Databases to Evaluate AIDS (CA-IeDEA) HIV policy model to inform equitable achievement of global goals, overall and across sub-populations, in Rwanda.

**Methods:** We created a deterministic dynamic model to project adult HIV epidemic and care continuum outcomes, overall and for 25 subpopulations (age group, sex, HIV acquisition risk, urbanicity). Data came from the Rwanda cohort of CA-IeDEA, 2004–2020; Rwanda Demographic and Health Surveys, 2005, 2010, 2015; Rwanda Population-based HIV Impact Assessment, 2019; and the literature and reports. We calibrated the model to 47 targets by selecting the 50 best-fitting parameter sets among 20,000 simulations. Calibration targets reflected epidemic (HIV prevalence, incidence), global goals (percentage on antiretroviral therapy (ART) among diagnosed, percentage virally suppressed among on ART) and other (number on ART, percentage virally suppressed) indicators, overall and by sex. Best-fitting sets minimized the summed absolute value of the percentage deviation (AVPD) between model projections and calibration targets. Good model performance was mean AVPD ≤5% across the 50 best-fitting sets and/or projections within the target confidence intervals; acceptable was mean AVPD >5% and ≤15%.

**Results:** Across indicators, 1,841 of 2,350 (78.3%) model projections were a good or acceptable fit to calibration targets. For HIV epidemic indicators, 256 of 300 (85.3%) projections were a good fit to targets, with the model performing better for women (83.3% a good fit) than for men (71.7% a good fit). For global goals indicators, 96 of 100 (96.0%) projections were a good fit; model performance was similar for women and men. For other indicators, 653 of 950 (68.7%) projections were a good or acceptable fit. Fit was better for women than for men (percentage virally suppressed only) and when restricting targets for number on ART to 2013 and beyond.

**Conclusions:** The CA-IeDEA HIV policy model fits historical data and can inform policy solutions for equitably achieving global goals to end the HIV epidemic in Rwanda. High-quality, unbiased population-based data, as well as novel approaches that account for calibration target quality, are critical to ongoing use of mathematical models for programmatic planning.

## INTRODUCTION

Rwanda, a Central African country with a generalized HIV epidemic (HIV prevalence = 3%) [1, 2], is accelerating toward achieving the 95-95-95 global targets and ending the public health threat posed by HIV epidemic by 2030 [3]. Recent data have generated optimism, with 97.5% of diagnosed Rwandan adults receiving antiretroviral therapy (ART) and 90.1% of Rwandan adults on ART having achieved HIV viral suppression [1]. However, Rwanda’s success is tempered by data suggesting lower HIV awareness (83.8% from population-based survey estimates); worse care continuum outcomes among men, adolescents and young adults; and notable HIV acquisition risk among rural populations [1, 4], which will impede continued progress and perpetuate existing disparities in achieving global targets.

Mathematical models are a tool commonly used to evaluate potential policy solutions addressing advancement toward epidemic control and health equity. For Rwanda, these models have been used to understand the distribution of new HIV infections [5]; evaluate the impact of HIV prevention policies on epidemic outcomes overall and among key population (i.e., female sex workers and their male clients) [6, 7]; forecast improvements along the care continuum on overall mortality among people with HIV [8]; and project achievement of behavioral and treatment targets on HIV transmission dynamics [9].

While the existing literature has used mathematical models primarily to assess the impact of different strategies on epidemic indicators, no mathematical models applied to Rwanda, to our knowledge, have examined potential policy solutions for achieving all global targets and promoting equity in achieving these targets across sub-populations. We aimed to develop and calibrate the Central Africa-International epidemiology Databases to Evaluate AIDS (CA-IeDEA) HIV policy model. IeDEA is an international research consortium that has collected and harmonized globally diverse observational data for over 2 million people living with HIV and enrolling in HIV care since 2004. The Rwanda cohort of CA-IeDEA comprises over 30,000 ever enrolled in HIV care at 12 clinics. It is thus a rich data source with which to populate mathematical models used to examine relevant, real-world policy solutions.

## MATERIALS AND METHODS

### Overview

We developed the CA-IeDEA HIV policy model, a deterministic dynamic compartmental model that projects HIV epidemic and care continuum outcomes. After model implementation, instantiation and parameterization, we calibrated the model to historical data, selected the best-fitting sets of model parameters, and evaluated the model’s performance. Model parameters and calibration targets were based on individual-level data from the Rwanda cohort of CA-IeDEA [10, 11], as well as Rwandan national surveys [12–14], international databases [15] and the literature (see Table 1). Secondary data were accessed 7 April 2022; the authors did not have access in the data to readily identifiable information that could identify individual participants. The model was implemented in R version 4.2.1 (R Core Team, 2022). Additional details are in the **Supplement**.

**Table 1.** Key model parameters and calibration targets.

|  | Baseline value(s)<sup>a</sup> | Data source(s) |
| --- | --- | --- |
| **Panel A. Parameters** |  |  |
| Model instantiation |  |  |
| Total population | 4,729,488 | World Bank [15] |
| Population size of female sex workers | 12,278 | UNAIDS [36] |
| HIV prevalence, by sub-population | 0.27%–63% | DHS Rwanda (2005) [12], BBSS (2010) [37] |
| Proportions along the care continuum, by sub-population and CD4 stratum<sup>a</sup> | 0–85% | CA-IeDEA (Rwanda, 2004) [11] |
| Force of infection |  |  |
| Probability of HIV transmission per sex act | 0.0005–0.0087 | Boily (2009) [39] |
| Proportion consistently using condoms | 4%–62% | DHS Rwanda (2005, 2010, 2015) [12–14], BBSS (2010) [37] |
| Effectiveness of condoms | 0.80 | Weller (2002) [40] |
| Effectiveness of viral suppression on HIV transmission | 0.966 | Supervie (2014) [41] |
| Number of sex acts | 1–40 | Braunstein (2011a) [42] |
| Transition probabilities (monthly) for infectious compartments |  |  |
| Susceptible population growth rates | 0.09%–0.45% | World Bank [15] |
| Natural history disease progression, by sex | 0.008–0.099 | CA-IeDEA (Rwanda, 2004–2020) [11] |
| Diagnosis, by CD4 stratum and year | 0.016–0.241 | DHS Rwanda (2005, 2010, 2015) [12–14], Okal (2017) [43] |
| Linkage to care, by CD4 stratum and year | 0.075–0.207 | Nsanzimana (2015) [44], Braunstein (2011b) [38], Okal (2017) [43] |
| Loss to follow-up, by CD4 stratum and ART status | 0.011–0.021 | CA-IeDEA (Rwanda, 2004–2020) [11] |
| On ART and virally suppressed, by CD4 stratum and year | 0.003–0.148 | CA-IeDEA (Rwanda, 2004–2020) [11] |
| ART failure, by CD4 stratum | 0.001–0.002 | CA-IeDEA (Rwanda, 2004–2020) [11] |
| Re-engagement in care<sup>b</sup> | 0.023 | CA-IeDEA (Rwanda, 2004–2020) [11] |
| Death on ART and virally suppressed, by sub-population and CD4 stratum<sup>b</sup> | 0.0002–0.017 | CA-IeDEA (Rwanda, 2004–2020) [11] |
| **Panel B. Calibration targets** |  |  |
| HIV epidemic |  |  |
| HIV prevalence (15–44 years), overall and by sex and year | 0.018–0.037 | DHS Rwanda (2005, 2010, 2015) [12–14], RPHIA (2019) [1] |
| HIV prevalence (15–64 years), overall and by sex | 0.022–0.037 | RPHIA (2019) [1] |
| HIV incidence (15–44 years), overall and by sex | 0.0006–0.0010 | RPHIA (2019) [1] |
| Global goals |  |  |
| % on ART among diagnosed (15–44 years), overall and by sex | 96.2%–97.2% | RPHIA (2019) [1] |
| % virally suppressed among on ART (15–44 years), overall and by sex | 75.9%–89.9% | RPHIA (2019) [1] |
| Other |  |  |
| % virally suppressed among diagnosed (15–44 years), overall and by sex | 61.3%–76.5% | RPHIA (2019) [1] |
| % virally suppressed among diagnosed (15–64 years), overall and by sex | 66.8%–77.2% | RPHIA (2019) [1] |
| Number on ART (15–44 years), overall | 6,356–197,544 | Rwanda National Report [45] |

Abbreviations: UNAIDS=Joint United Nations Programme on HIV/AIDS; DHS=Demographic and Health Survey; BBSS=Behavioral and Biological Surveillance Survey; CA-IeDEA= Central Africa International epidemiology Databases to Evaluate AIDS; RPHIA= Rwanda Population-Based HIV Impact Assessment.
<sup>a</sup> The baseline value(s) reporting a range represent the range of baseline parameter values or calibration targets across all sub-populations and years. The parameter values and calibration targets, overall and for each sub-population, are reported separately in the supplementary material.

### CA-IeDEA HIV policy model

#### Overall structure

The CA-IeDEA HIV policy model is an extended Susceptible-Infected-Removed (SIR) dynamic epidemic model, adapted from previous work [16]. The model includes a susceptible compartment, 24 mutually exclusive infectious compartments and a death compartment (). The susceptible compartment includes the population at risk of acquiring HIV. Infectious compartments capture HIV disease progression and steps along the HIV care continuum. We characterize HIV disease progression according to different CD4 cell count strata (>500, 351–500, 201–350 and <200 cells/mm^3^) that reflect key thresholds for the risk of HIV-related opportunistic infections and other illnesses [17]. They also correspond to international and Rwandan recommendations on CD4-based thresholds for antiretroviral therapy (ART) initiation over time [18–24]. We did not explicitly model acute HIV, given its small window of detection (approximately 14 days) that is captured in a single model time step (see below).

The HIV care continuum is defined as key steps along the care cascade: undiagnosed, diagnosed, linked to care, on ART and virally suppressed, on ART and not virally suppressed, and lost to follow-up. We explicitly modeled linkage to care due to historical delays in ART initiation [25, 26], including rapid ART initiation (e.g., within 7 days), after linkage to care [21, 25, 26]. We did not explicitly model on-ART disease progression to maintain model parsimony and given successful calibration of other similarly structured HIV policy models [16, 27]. Death occurs from background mortality and HIV-related causes, with transitions to the death compartment occurring only from infectious compartments. There is no transition from the susceptible to the death compartment as changes in the size of the susceptible population over time reflect background mortality.

Model dynamics are expressed using a system of ordinary differential equations. The differential equations are implemented as difference equations in order to project outcomes at discrete time points [28], which is important for informing policy decisions, and also to reduce computational complexity during the model calibration process. Movement between compartments occurs based on a one-month time step, which captures clinically meaningful events related to disease progression and engagement in care.

#### Study population partitions

The model is partitioned into 25 sub-populations by age, sex, risk and subnational unit (i.e., urbanicity) to account for differences in HIV acquisition risk and steps along the care continuum, resulting in a total of 650 model compartments (25 sub-populations x 26 compartments). Age is partitioned into five groups (i.e., 15–24, 25–34, 35–44, 45–54, 55+ years) given age-related differences in HIV prevalence and HIV testing [29–32]. Women are represented as having either lower or higher HIV acquisition risk, with the latter representing female sex workers (FSWs) who have higher HIV prevalence (45.8% compared to 3.6% among low-risk women) and lower viral suppression (71% compared to 79% among lower-risk women) [33]. Men are considered lower risk. We do not explicitly include male clients of female sex workers as a higher risk sub-population, given the HIV acquisition risk for male clients of female sex workers is similar to non-clients [29–31]. Subnational units are applied as an urban-rural classification because HIV prevalence in urban areas in Rwanda is more than double that of rural areas [21]. However, we did not partition women who were at higher risk of HIV acquisition (i.e., FSWs) by urbanicity given more than 80% reside in urban areas [34].

#### Force of infection

The force of infection, or the probability of HIV acquisition in different susceptible sub-populations, is based on heterosexual contact. We calculate the force of infection using: the per-act HIV transmission probability in the absence of condoms and HIV treatment, the probability of condom use, the probability of having a virally suppressed partner, the reduction in HIV acquisition risk due to condom use or having a virally suppressed partner, the probability of sexual contact with a specific sub-population, the probability of a sexual partner with HIV within a sub-population, and the number of sex acts per month. We assume no sexual contact between lower risk populations living in urban and in rural areas due to relatively low spatial mobility in Rwanda [35]; however, FSWs have sexual contact with sub-populations living in either urban or rural areas. We apply random mixing across sub-populations with sexual contacts given our assumption that all sub-populations are sexually active and randomly choose their sexual partners, including FSWs.

### Model inputs

#### Instantiation

Model instantiation involved distributing the total population across sub-populations and by HIV status, disease progression and care engagement (**Table 1**). In the current study, the initial distribution was applied to the overall Rwandan adult population 15–64 years in 2004, the first year in the historical projection period used for the calibration process. We determined the total number in each sub-population based on population projections and the literature specific to Rwanda [15, 36], estimating the number of people living with HIV in a given sub-population from national survey data on HIV prevalence (Demographic and Health Survey (DHS) Rwanda, 2005; Behavioral and Biological Surveillance Survey (BBSS), 2010) [12, 37].

For each infectious compartment of a given sub-population, the initial population was calculated based on the proportion living with HIV in each step of the HIV care continuum and in each CD4 stratum. These proportions were estimated using individual-level longitudinal data on HIV treatment and care from the Rwanda cohort of CA-IeDEA [11], as well as national surveys (DHS Rwanda 2005) [12] and the literature [38].

#### Parameterization

Model parameterization involved deriving parameters, by sub-population, for susceptible population growth, force of infection, and monthly transition probabilities between compartments (**Table 1**). We assumed that the monthly population growth rate applied to the susceptible compartments was equal to the total population growth rate, which we estimated as the simple average of the Rwanda’s monthly population growth rate, 2004–2020, from World Bank population projections [15]. Parameter values for the force of infection were generally extracted from literature synthesizing existing evidence from sub-Saharan African countries or surveys conducted in Rwanda, with the proportion using condoms estimated directly from DHS Rwanda (2005, 2010, 2015) for the lower risk population and from the BBSS report for FSWs [12–14, 37].

For all sub-populations except FSWs, transition probabilities for natural history disease progression, HIV viral suppression, ART failure, loss to follow-up, care re-engagement and death were derived from the Rwanda cohort of CA-IeDEA, 2004–2020 [11], while those for HIV diagnosis and linkage to care were estimated from DHS Rwanda (2005, 2010, 2015) and extracted from the literature [12–14, 44], respectively. We defined HIV viral suppression as individuals on ART without evidence of virologic failure, which was at least one post-ART initiation follow-up HIV viral load test result ≥1000 copies/mL [24]. We defined loss to follow-up as at least 365 days between the last clinic visit and last contact (among those on ART) or at least 6 months between the most recent clinic visit and last contact (pre-ART) [43, 46]. Monthly probabilities of death accounted for misclassification of loss to follow-up both on and off ART [47, 48].

For FSWs, transition probabilities for diagnosis, linkage to care and condom use were extracted from existing literature and government reports [38, 39, 41]. When transition probabilities for loss to follow-up and re-engagement in this sub-population were unavailable, we applied the baseline values used for the lower risk female population but applied wider confidence bounds to account for the uncertainty in the parameter value.

### Model calibration

We calibrated model parameter values in order to fit model projections to historical data. The process involved identifying calibration targets, generating and selecting best-fitting sets of model parameters, and assessing the model’s performance.

#### Calibration targets

We selected 47 calibration targets reflecting HIV epidemic, global goals and other care engagement indicators for adults 15–44 years and 15–64 years, overall and/or by sex, over a 16-year historical calibration period between 2004 and 2020 (Table 1). HIV epidemic targets included HIV prevalence and HIV incidence and came from DHS (Rwanda, 2005, 2010, 2015) and RPHIA (2019) [1, 29–31]. Global goals targets included the percentage on ART among adults diagnosed with HIV and percentage virally suppressed among adults on ART (RPHIA, 2019) [1]. We did not include global goals targets for youth 15–24 years due to small sample sizes in available survey data. Finally, other targets included the percentage virally suppressed among adults living with HIV (RPHIA, 2019) and the number of adults on ART (Rwanda National HIV Report, 2020) [1, 45]. Because available data report all Rwandans on ART, including pediatric cases, we estimated the number of adults on ART as the product of the total reported number on ART and the proportion reported on ART within a given age group (e.g., 15–44 years) among total population on ART [45].

#### Selecting best-fitting sets of model parameters

To select best-fitting parameter sets, we first generated sets of model parameters. We assigned distributions to all model parameters, with the exception of the population size for each sub-population and the proportions of people living with HIV by CD4 stratum given these parameters are single point estimates. We assigned the normal distribution to the susceptible population growth rate; beta distributions to transition probabilities, except the probabilities of linkage to care due to limited information on sample size; and triangular distributions to model parameters that lacked information on their distribution. Across relevant parameters, we then performed a random draw of each parameter’s value from its distribution in order to generate a parameter set. We applied random draws to the over 1,500 model parameters and generated 20,000 randomly selected parameter sets to fully capture variability across model parameters. Each parameter set was then used to project historical model outcomes associated with the set.

We selected the best-fitting sets of model parameters by examining their goodness of fit (GoF), a measure quantifying the difference between the model’s projected outcomes and calibration targets. No standard GoF measure(s) exists and thus GoF is defined according to a given model structure and selected calibration targets [49]. In the current analysis, the GoF measure was defined as the summed absolute value of the percentage deviation (sAVPD) between the model’s projected outcomes and the calibration targets, with the sAVPD a scale-independent measure selected to standardize differences in the units of analysis across calibration targets [50, 51]. A smaller sAVPD value represents a smaller overall difference between model projections and the historical data, indicating a better model fit. The 50 sets that minimized the sAVPD were considered the best-fitting sets of model parameters.

#### Performance of the calibrated model

Model performance was evaluated according to the percentage of model projections considered good or acceptable. ‘Good’ model performance was a mean AVPD ≤5% between the model’s projected outcomes and the calibration target(s) across the top 50 best-fitting parameter sets, while ‘acceptable’ performance was defined as a mean AVPD >5% and ≤15% [52, 53]. If a model projection was with a given target’s confidence intervals, performance was also considered ‘good’ regardless of the mean AVPD. Among model projection(s) considered ‘good’, visual trend(s) were required to reflect trends in the historical data, although this was a subjective assessment and only occurred if sufficient targets were available. Model performance was evaluated separately for HIV epidemic indicators, global goals indicators and other indicators.

## RESULTS

### HIV epidemic indicators

For the overall adult population (15–44 and 15–64 years), we compared 300 model projections for HIV epidemic indicators to the calibration targets (50 projections/target x 6 targets=300). Among these 300, 256 (85.3%) were a good fit to calibration targets and 23 (7.6%) an acceptable fit (**Fig 2)**. For HIV prevalence indicators only (5 targets), 216 (86.4%) projections were a good fit to calibration targets and 23 (9.2%) were an acceptable fit. Model performance worsened when fitting to targets in 2019, with at least 47 (94.0%) projections a good fit for 2005, 2010 and 2015 targets, but less than 32 (64.0%) projections a good fit to 2019 targets. For the HIV incidence indicator only (1 target), 50 (100%) projections were good fit to the calibration targets; assessing trends over time was not possible given only a single target.

**Fig 1.**
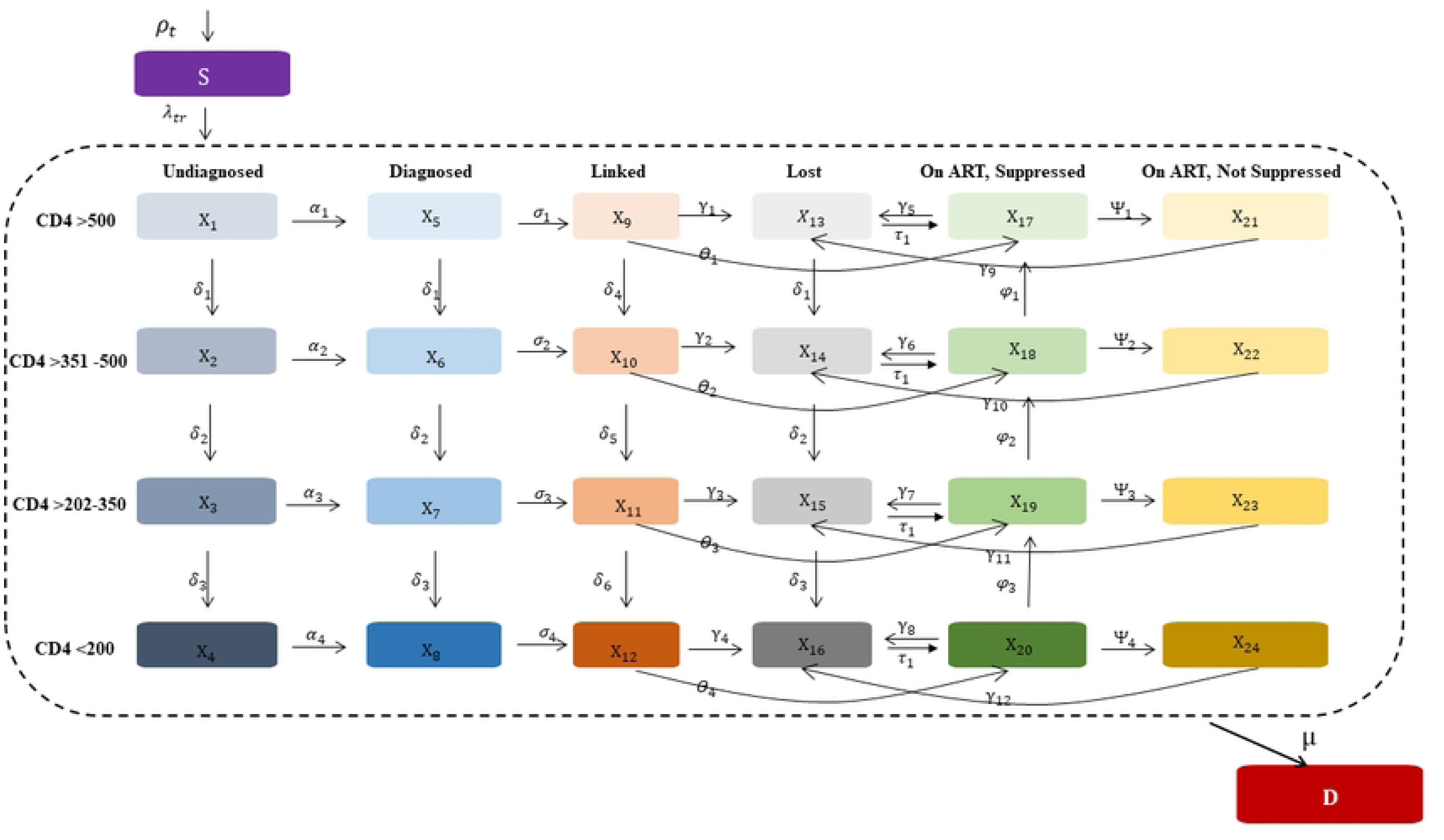
Model schematic of the CA-IeDEA HIV policy model. *S=Susceptible compartment; I=Infectious compartments; D=Death compartments; ART=antiretroviral.* Shown is a simplified schematic of the CA-IeDEA HIV policy model. Each modeled population enters the susceptible compartment, can probabilistically transition to and move between infectious compartments and can probabilistically transition to the death compartment, with transitions and their directionality denoted by arrows. Infectious model compartments are outlined by the dotted line and represent the combination of degree of HIV disease progression (i.e., CD4 cell count strata) and engagement in HIV care; disease progression for those on ART represents the CD4 cell count stratum at ART initiation. Each model compartment includes 25 sub-populations partitioned by age, sex, risk of acquisition and urbanicity (sub-populations not shown).

**Fig 2.**
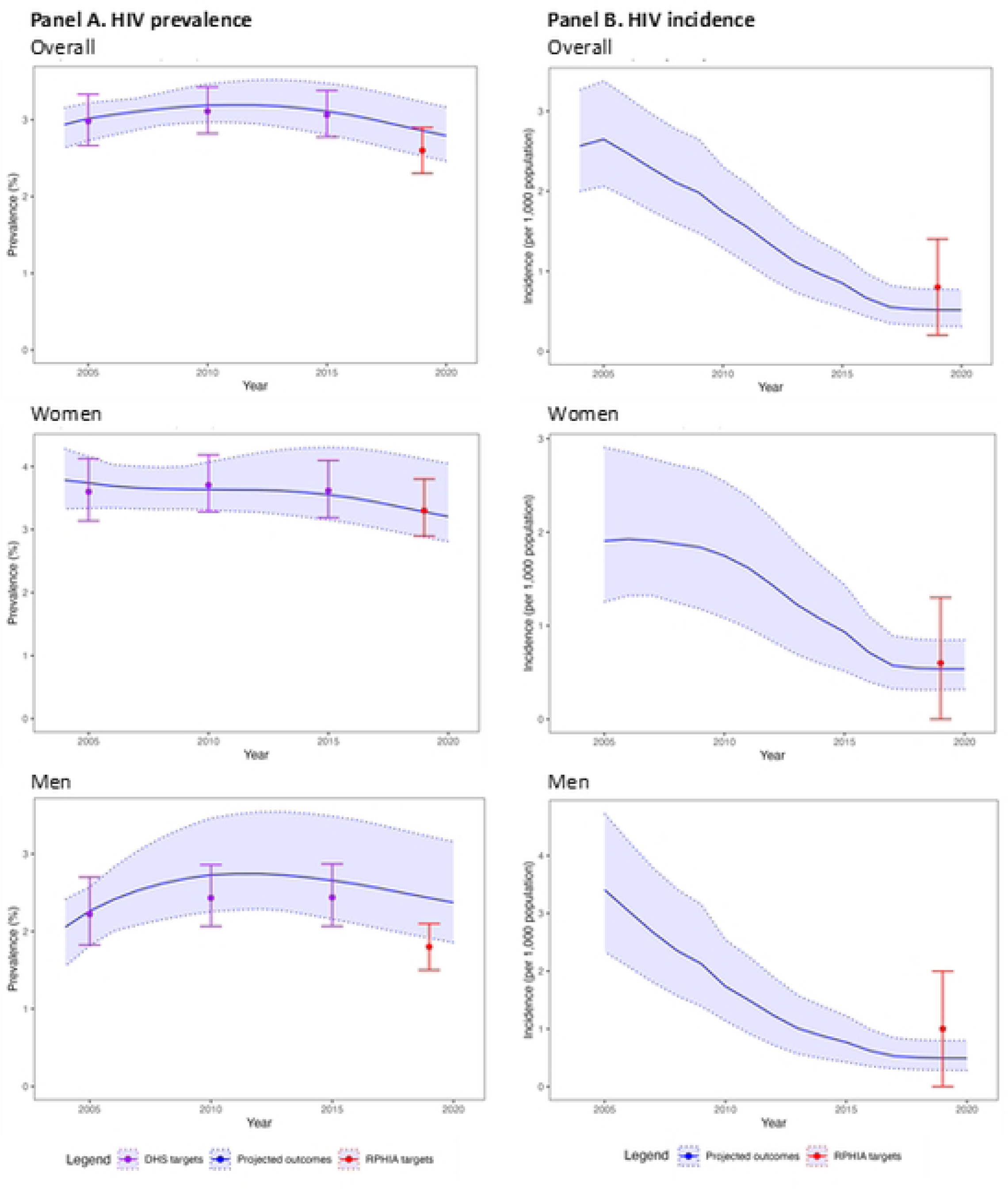
Model projections and HIV epidemic calibration targets for 15–44 year-olds in Rwanda, 2004–2020. *Abbreviation: DHS=Demographic Health Survey; RPHIA=Rwanda Population-based Health Impact Assessment* The blue line and light blue shades represent the mean and range of the model projections, respectively, using the 50 best-fitting parameter sets. The points and the error bar represent the point estimates and 95% confidence interval of the calibration targets, with targets from DHS Rwanda data in purple and targets from RPHIA in red. (A) For HIV prevalence indicators, 80.9% of model projections have good or acceptable fit to calibration targets, with model performance better for women than for men particularly later in the calibration period. (B) For HIV incidence, 100% of model projections have a good fit, which is consistent for women and men.

For the adult female and male sub-populations (15–44 and 15–64 years), we examined 300 model projections each for women and for men (50 projections/target x 6 targets=300 for each sub-population). The model performed better for women than for men, with 250 (83.3%) and 215 (71.7%) projections having a good fit, respectively (Fig 2). An additional thirteen projections for women were an acceptable fit, but none for men. For HIV prevalence indicators only, 200 (80.0%) projections for women and 165 (66.0%) for men were a good fit. Model performance was consistently good (≥96% good fit) for women 15-44 years over time; however, performance varied for men in this age group, with over 80% of projections a good fit in 2005 and 2015, but only 8 (16.0%) a good fit in 2019. In contrast, among the population age 15–64 years, the model performed better for men (68.0% of projections a good fit) compared to women (10.0% of projections a good fit and 22% an acceptable fit). For the HIV incidence indicator, 50 (100%) projections were a good fit to calibration targets for both women and men.

### Global goals indicators

For the overall adult population (15–44 years), we examined 100 model projections for global goals indicators (50 projections/target x 2 targets=100) (**Fig 3, Panels A and B**). Among these 100, 96 (96.0%) projections were a good fit and 4 (4.0%) were acceptable. For the percentage on ART among adults diagnosed with HIV (1 target), 50 (100.0%) projections were a good fit to calibration targets. Visually, the projected percentages decreased in both 2005 and 2011, but had sharp increases to reflect trend in the subsequent projection periods. For percentage virally suppressed among adults on ART (1 target), 46 (92.0%) projections were good fit to targets with an additional 4 (8.0%) projections being acceptable.

**Fig 3.**
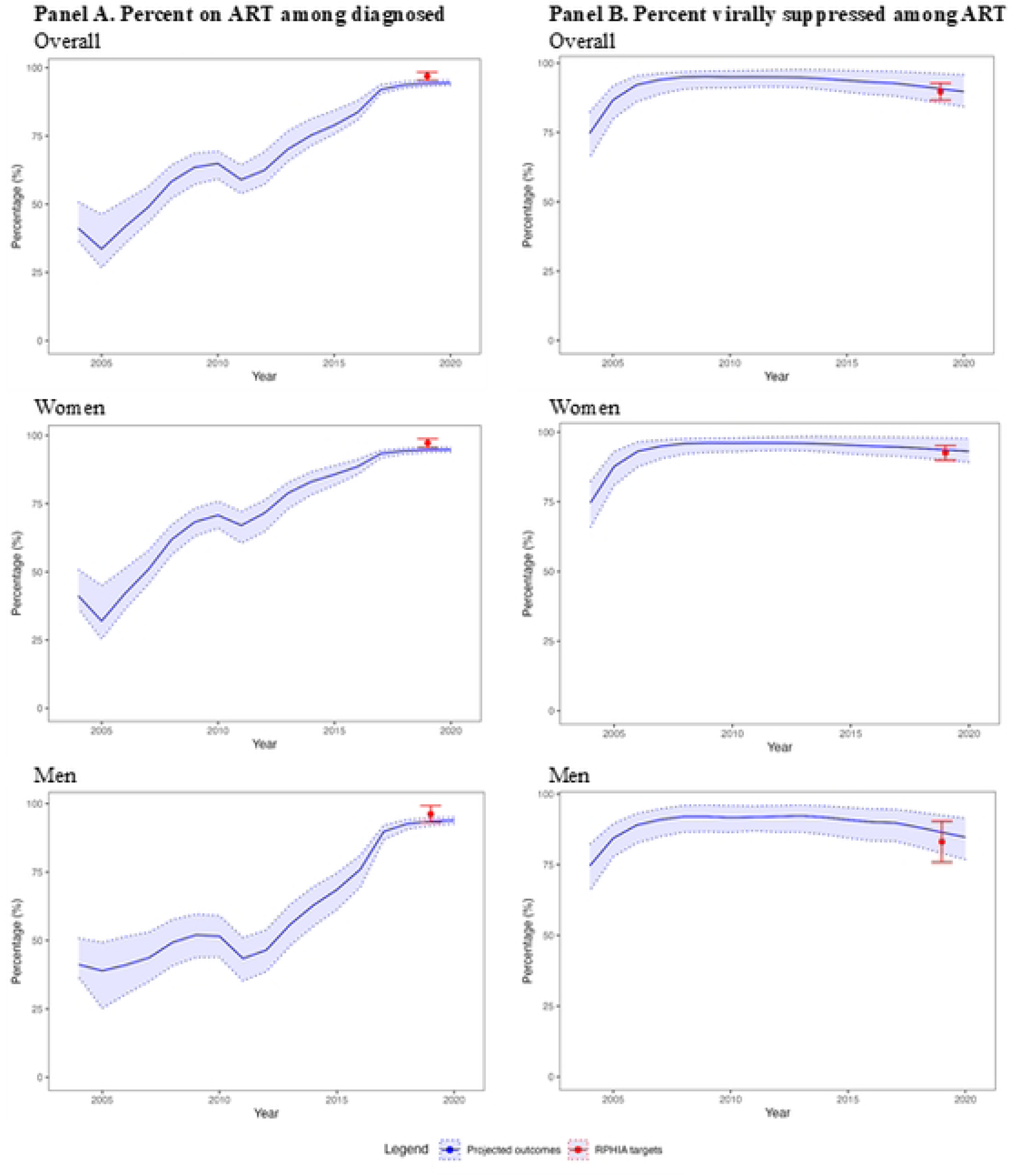
Model projections and global goals calibration targets for 15–44 year-olds in Rwanda, 2004–2020. *Abbreviation: DHS=Demographic Health Survey; RPHIA=Rwanda Population-based Health Impact Assessment* The blue line and light blue shades represent the mean and range of the model projections, respectively, using the 50 best-fitting parameter sets. The points and the error bar represent the point estimates and 95% confidence interval of the calibration targets, with targets from DHS Rwanda data in purple and targets from RPHIA in red. (A) For the percentage on ART among diagnosed adults, 100% of model projections had good fit to calibration targets. (B) For the percent virally suppressed among adults on ART, 92% of model projections had a good fit. (C) For the number of adults on ART, 28% of model projections had a good fit.

For the adult female and male sub-populations (15–44 years), we examined 100 projections each for women and for men (50 projections/target x 2 targets=100 for each sub-population). Here, 98 (98.0%) and 94 (94.0%) projections were good fit for women and men, respectively. Two additional projections were acceptable for women and six for men. For percentage on ART among adults diagnosed with HIV, 50 (100%) projections were good fit to calibration targets for both women and men. For percentage virally suppressed among adults on ART, model performance is slightly better for women than men, with 48 (96.0%) and 44 (88.0%) being good fit to calibration targets, respectively.

### Other indicators

For the overall adult population (15–44 and 15–64 years), we examined projections for the number on ART (50 projections/target x 18 targets=850) and the percentage virally suppressed among the diagnosed adult population with HIV (50 projections/target x 2 targets=100). For number on ART, 573 (67.4%) projections were good or acceptable fit with two-fifths of these projections a good fit (**Fig 4**). Model fit for the number on ART improved when restricting the targets to years 2013 and beyond (8 targets), where 369 (92.3%) were good or acceptable. For the percentage virally suppressed, 80 (80.0%) projections had a good or acceptable fit with less than one-fifth of these projections considered a good fit. Here, the model performed better among adults age 15–64 years (94.0% of projections a good or acceptable fit) than adults age 15–44 years (67.0% of projections a good or acceptable fit).

**Fig 4.**
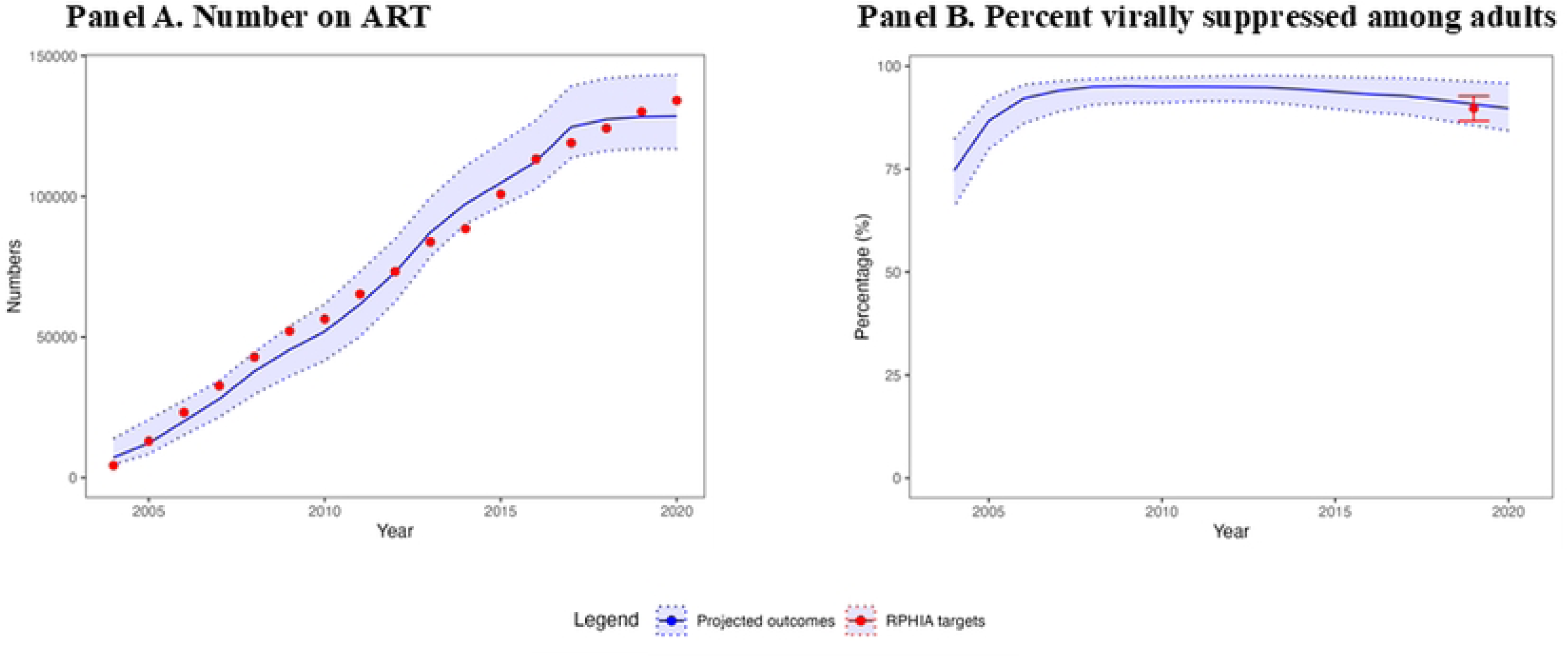
Model projections and other calibration targets for 15–44 year-olds in Rwanda, 2004–2020. *Abbreviation: RPHIA=Rwanda Population-based Health Impact Assessment* The blue line and light blue shades represent the mean and range of the model projections, respectively, using the 50 best-fitting parameter sets. The points and the error bar represent the point estimates and 95% confidence interval of the calibration targets, with targets from DHS Rwanda data in purple and targets from RPHIA in red (A) For the number of adults on ART, 28% of model projections had a good fit. (B) For the percentage virally suppressed among adults living with HIV, 65% of model projections had good or acceptable fit to calibration targets.

For the adult female and male sub-populations (15–44 and 15–64 years), we examined 100 model projections each for women and men on percentage virally suppressed among people with HIV (50 projections/target x 2 targets=100 for each sub-population). Here, 77 (77%) projections for women and 54 (54%) for men had good or acceptable fit. For both age ranges, a smaller proportion of projections had a good fit for women (less than 5%) than for men (20%– 40%). Model fit for number on ART was not assessed by sub-population due to lack of data.

## DISCUSSION

Leveraging the Rwanda cohort of CA-IeDEA and other nationally representative data, we developed and calibrated the CA-IeDEA HIV policy model that will inform policy solutions for equitably achieving global targets. Based on 50 best-fitting sets of model parameters, we found that the policy model has a good fit to HIV epidemic and global goals (i.e., 95-95-95) targets over the calibration period, with better performance for women than for men for most targets. Model performance was acceptable for the number on ART in recent years and percentage virally suppressed among adults 15–64 years.

Across HIV epidemic and global goals targets, we found that model projections for women outperformed those for men in most cases. This could be explained by greater uncertainty in model parameters for men than for women [54, 55]. For example, among men, model projections for HIV incidence were consistently towards the lower bound of the target confidence intervals, but increasingly overestimated HIV prevalence over the calibration period. This could result from model parameters that overestimate the proportion of condom use among men, which results in underestimation of the probability of HIV transmission, lower projected HIV incidence and higher HIV prevalence. In our model, the parameters for the proportion of men using condoms were based on self-reported condom use [29–31] . Self-reported condom use may be affected by social desirability bias, a type of response bias suggesting respondents may answer questions in a way that is likely to be viewed more favorably [56]. In this case, the socially desirable response is to report consistent condom use with sexual partners. Men have been found to consistently report higher levels of condom use compared to women’s reporting of condom use by their male partners [57], suggesting overreporting among men. Improved data quality can reduce uncertainty in parameters and improve model performance.

For HIV epidemic indicators, namely HIV prevalence, we found that model fit later in the calibration period (2019) was not as good as earlier in the calibration period (2005–2015). A possible explanation is uncertainty in HIV prevalence targets because they come from two independent data sources, variation in the number of targets available for a given data source and differences in methodologic approach across sources. That is, three targets came from DHS (2005, 2010, 2015), while one target came from PHIA (2019). This resulted in higher weight of DHS targets during the model calibration process. This resultant weighting is notable because the DHS and PHIA data used different sample selection approaches to estimate HIV prevalence. DHS data estimated HIV prevalence among Rwandans who voluntarily received anonymous HIV testing (i.e., opt-in approach) [31]; in contrast, PHIA data estimated HIV prevalence among individuals who did not refuse to receive their HIV testing result (i.e., opt-out approach) [1]. Evidence suggests that opt-out HIV testing has a higher uptake than opt-in testing [58], which implies a less biased sample for PHIA estimates. Ultimately, the estimate from PHIA is less likely to follow the model’s HIV prevalence trend fitted to DHS data. Future work to consider data quality in model calibration targets—an area understudied in the literature—is warranted.

While the model generally had a good fit to HIV epidemic and global goals calibration targets, the model did not perform as well for other targets. For percentage virally suppressed among diagnosed adults, for example, a possible explanation is potential bias in the targets. That is, the targets were calculated and reported conditional on self-reported awareness of HIV-positive status. However, evidence suggests that underreporting of awareness of HIV-positive status in observational data occurs consistently across countries, ranging from 3% (Eswatini) to 47% (Nigeria) [59–62]. Our model’s worse fit to these targets may reflect potential biases in the data or, alternatively, may reveal plausible trends in care engagement in this setting.

Model fit for number on ART, 15–44 years, similarly did not perform as well as for HIV epidemic or global goals indicators, although fit was better later in the calibration period. Worse performance could be due in part to the aggregated data available for this calibration target: That is, available data report all Rwandans on ART, including pediatric cases. Although we adjusted the targets based on the age distribution of Rwandans on ART, the age distribution we applied was available only in the most recent data year for this target (2021) [63]. Model fit could be improved with additional data that disaggregate the information by age and time.

The CA-IeDEA HIV policy model, designed explicitly to assess potential policy solutions for equitably achieving global targets in Rwanda, extends existing mathematical models applied in the Rwandan context. Our model includes two broad dimensions (disease progression and the HIV care continuum) versus only a single dimension (care continuum) as in the Nsanzimana model [6]. Including these two dimensions better account for differences in health policy (e.g., ART initiation criteria) over time, improved model fit to historical data and increased potential to capture more nuanced differences in future health outcomes overall and for different sub-populations. Further, our model included 25 sub-populations defined by age, gender, risk and sub-national unit versus risk only (as in the Nsanzimana model) or age and gender only (as in the Bendavid model) [6, 8]. The CA-IeDEA HIV policy model’s structure will allow for more nuanced assessment of programmatic needs to achieve country-specific goals and do so equitably across sub-populations [31].

The CA-IeDEA HIV policy model also adds to existing mathematical modeling literature methodologically. Systematic reviews find that a large proportion of modeling studies do not specify quantitative GoF measures for model calibration [64, 65]. Indeed, the two existing modeling studies in Rwanda provide no description of GoF measures. While our approach, percent deviation, has been used less commonly [65], it aligns with modeling studies that use calibration targets with disparate scales [50]. That is, a percent deviation GoF measure allows for inclusion of a variety of calibration targets and from multiple data sources, as in Rwanda, in order to improve model performance [44].

### Limitations

Our model is not without limitations. First, we did not explicitly include men who have sex with men (MSM) as a sub-population in our model, although model revisions to include this sub-population are in development. Second, we assumed no urban-rural sexual contacts. While this may lead to underestimation of HIV incidence over time due to urbanization and increased mobility in Rwanda [66], the model had a good fit to HIV incidence indicators over the calibration period. Third, in deriving model parameters, we did not explicitly account for potential bias inherent in the observational data. However, in the calibration process, we addressed this limitation by allowing for full variation in the baseline parameter values derived from the observational data. Fourth, we did not specifically capture either in the model or in the model parameters social and health system contexts (e.g., women empowerment, trust in HIV services) in Rwanda [67], which might limit the accuracy of model projections. However, by allowing for variation in parameter values, we likely account for the effects of social and health system factors on HIV epidemic, global goals and other indicators.

Fifth, calibration targets for certain sub-populations (e.g., adolescent girls and young women, female sex workers) were limited given data unavailability. However, model fit can be re-assessed as population-based data emerge for these sub-populations. Lastly, our GoF measure, i.e., percent deviation, may overweight calibration targets with small baseline values because small differences between projected and calibrated values can translate into a large percentage differences. Similarly, our GoF measure did not capture differences in the quality of calibration target data. To minimize such effects, we integrated an additional model performance measure on the percentage of model projections within calibration target CIs, when available.

## CONCLUSIONS

The calibrated CA-IeDEA HIV policy model is ideally positioned to examine the policy solutions that will propel Rwanda not only toward ending the HIV epidemic as a public health threat, but doing so equitably. As mathematical modeling continues to be used to inform programmatic planning, there is a critical need for high quality population-based calibration target data over time, as well as development of approaches to measure and adjust for calibration target quality and refine GoF measures when relying on disparate scales.

## Data Availability

All relevant data are within the manuscript and its Supporting Information files.

## ACKNOWLEDGEMENTS

Members of Central Africa IeDEA are: Nimbona Pélagie, Association Nationale de Soutien aux Séropositifs et Malade du Sida (ANSS), Burundi; Patrick Gateretse, Jeanine Munezero, Valentin Nitereka, Annabelle Niyongabo, Zacharie Ndizeye, Christella Twizere, Théodore Niyongabo, Centre National de Référence en Matière de VIH/SIDA, Burundi; Hélène Bukuru, Thierry Nahimana, Martin Manirakiza,Centre de Prise en Charge Ambulatoire et Multidisciplinaire des PVVIH/SIDA du Centre Hospitalo-Universitaire de Kamenge (CPAMP-CHUK), Burundi; Patrice Barasukana, Hélène Bukuru, Martin Manirakiza, Zacharie Ndizeye, CHUK/Burundi National University, Burundi; Jérémie Biziragusenyuka, Ella Ange Kazigamwa, Centre de Prise en Charge Ambulatoire et Multidisciplinaire des PVVIH/SIDA de l’Hôpital Prince Régent Charles (CPAMP-HPRC), Burundi; Caroline Akoko, Ernestine Kesah, Esther Neba, Denis Nsame, Vera Veyieeneneng, Bamenda Regional Hospital, Cameroon; Bazil Ageh Ajeh, Rogers Ajeh, Dan Ebai Ashu, Eta Atangba, Christelle Tayomnou Deussom, Peter Vanes Ebasone, Ernestine Kendowo, Clarisse Lengouh, Gabriel Mabou, Sandra Mimou Mbunguet, Judith Nasah, Nicoline Ndiforkwah, Marc Lionel Ngamani, Eric Ngassam, George Njie Ngeke, Clenise Ngwa, Anyangwa Sidonie, Clinical Research Education and Consultancy (CRENC), Cameroon; Anastase Dzudie, CRENC and Douala General Hospital, Cameroon; Djenabou Amadou, Joseph Mendimi Nkodo, Eric Pefura Yone, Jamot Hospital, Cameroon; Annereke Nyenti, Phyllis Fon, Mercy Ndobe, Priscilia Enow, Limbe Regional Hospital, Cameroon; Catherine Akele, Akili Clever, Faustin Kitetele, Patricia Lelo, Kalembelembe Pediatric Hospital, Democratic Republic of Congo; Nana Mbonze, Guy Koba, Martine Tabala, Cherubin Ekembe, Didine Kaba, Kinshasa School of Public Health, Democratic Republic of Congo; Jean Paul Nzungani, Simon Kombela, Dany Lukeba, Sangos plus/Bomoi, Democratic Republic of Congo; Mattieu Musiku, Clement Kabambayi, Job Nsoki,Hopital de Kabinda, Democratic Republic of Congo; Merlin Diafouka, Martin Herbas Ekat, Dominique Mahambou Nsonde, CTA Brazzaville, Republic of Congo; Ursula Koukha, Adolphe Mafoua, Massamba Ndala Christ, CTA Pointe-Noire, Republic of Congo; Jules Igirimbabazi, Nicole Ayinkamiye, Bethsaida Health Center, Rwanda; Providance Uwineza, Emmanuel Ndamijimana, Busanza Health Center, Rwanda; Jean Marie Vianney Barinda, Marie Louise Nyiraneza, Gahanga Health Center, Rwanda; Marie Louise Nyiransabimana, Liliane Tuyisenge, Gikondo Health Center, Rwanda; Catherine Kankindi, Christian Shyaka, Kabuga Health Center, Rwanda; Bonheur Uwakijijwe, Marie Grace Ingabire, Kicukiro Health Center, Rwanda; Beltirde Uwamariya, Jules Ndumuhire, Masaka Health Center, Rwanda; Gerard Bunani, Fred Muyango, Nyagasambu Health Center, Rwanda; Yvette Ndoli, Oliver Uwamahoro, Nyarugunga Health Center, Rwanda; Eugenie Mukashyaka, Rosine Feza, Shyorongi Health Center, Rwanda; Chantal Benekigeri, Jacqueline Musaninyange, WE-ACTx for Hope Clinic, Rwanda; Josephine Gasana, Charles Ingabire, Jocelyne Ingabire, Faustin Kanyabwisha, Gallican Kubwimana, Fabiola Mabano, Jean Paul Mivumbi, Benjamin Muhoza, Athanase Munyaneza, Gad Murenzi, Francoise Musabyimana, Allelluia Giovanni Ndabakuranye, Fabienne Shumbusho, Patrick Tuyisenge, Francine Umwiza, Research for Development (RD Rwanda) and Rwanda Military Hospital, Rwanda; Jules Kabahizi, Janviere Mutamuliza, Boniface Nsengiyumva, Ephrem Rurangwa, Rwanda Military Hospital, Rwanda; Eric Remera, Gallican Nshogoza Rwibasira, Rwanda Biomedical Center, Rwanda. Adebola Adedimeji, Kathryn Anastos, Jean Claude Dusingize, Lynn Murchison, Viraj Patel, Jonathan Ross, Marcel Yotebieng, Natalie Zotova, Albert Einstein College of Medicine, USA; Ryan Barthel, Ellen Brazier, Heidi Jones, Elizabeth Kelvin, Denis Nash, Saba Qasmieh, Chloe Teasdale, Institute for Implementation Science in Population Health, Graduate School of Public Health and Health Policy, City University of New York (CUNY), USA; Batya Elul, Columbia University, USA; Xiatao Cai, Don Hoover, Hae-Young Kim, Chunshan Li, Qiuhu Shi, Data Solutions, USA; Kathryn Lancaster, The Ohio State University, USA; Mark Kuniholm, University at Albany, State University of New York, USA; Andrew Edmonds, Angela Parcesepe, Jess Edwards, University of North Carolina at Chapel Hill, USA; Olivia Keiser, University of Geneva; Stephany Duda; Vanderbilt University School of Medicine, USA; April Kimmel, Virginia Commonwealth University School of Medicine, USA.

